# Evaluating the accuracy and reliability of large language models in assisting with pediatric differential diagnoses: A multicenter diagnostic study

**DOI:** 10.1101/2024.08.09.24311777

**Authors:** Masab A. Mansoor, Andrew F. Ibrahim, David J. Grindem, Asad Baig

**Affiliations:** Edward Via College of Osteopathic Medicine – Louisiana Campus; Texas Tech University Health Science Center; Mayo Clinic; Columbia University

**Keywords:** GPT-3, newer technology in healthcare, pediatrics, artificial intelligence in medicine, large language models

## Abstract

**Importance:** Large language models, such as GPT-3, have shown potential in assisting with clinical decision-making, but their accuracy and reliability in pediatric differential diagnosis in rural healthcare settings remain underexplored.

**Objective:** Evaluate the performance of a fine-tuned GPT-3 model in assisting with pediatric differential diagnosis in rural healthcare settings and compare its accuracy to human physicians.

**Methods:** Retrospective cohort study using data from a multicenter rural pediatric healthcare organization in Central Louisiana serving approximately 15,000 patients. Data from 500 pediatric patient encounters (age range: 0-18 years) between March 2023 and January 2024 were collected and split into training (70%, n=350) and testing (30%, n=150) sets.

**Interventions:** GPT-3 model (DaVinci version) fine-tuned using OpenAI API on training data for ten epochs.

**Main Outcomes and Measures:** Accuracy of fine-tuned GPT-3 model in generating differential diagnoses, evaluated using sensitivity, specificity, precision, F1 score, and overall accuracy. The model’s performance was compared to human physicians on the testing set.

**Results:** The fine-tuned GPT-3 model achieved an accuracy of 87% (131/150) on the testing set, with a sensitivity of 85%, specificity of 90%, precision of 88%, and F1 score of 0.87. The model’s performance was comparable to human physicians (accuracy 91%; P = .47).

**Conclusions and Relevance:** The fine-tuned GPT-3 model demonstrated high accuracy and reliability in assisting with pediatric differential diagnosis, with performance comparable to human physicians. Large language models could be valuable tools for supporting clinical decision-making in resource-constrained environments. Further research should explore implementation in various clinical workflows.

## Introduction

The rapid advancement of artificial intelligence (AI) has led to the development of large language models (LLMs) that have demonstrated remarkable capabilities in understanding, generating, and analyzing human language [1]. LLMs, such as GPT-3, have shown potential in various domains, including healthcare, where they can assist with tasks such as clinical decision support, patient engagement, and medical research [2-3]. In particular, LLMs have been explored for their ability to aid in diagnostic processes, such as generating differential diagnoses based on patient symptoms and medical history [4-5].

Differential diagnosis, distinguishing a particular disease or condition from others with similar clinical features, is a critical skill for healthcare providers [6]. In pediatric care, differential diagnosis can be particularly challenging due to the wide range of conditions that can present overlapping symptoms and the difficulty in obtaining accurate patient histories from young children [7]. Misdiagnosis or delayed diagnosis can lead to inappropriate treatment, prolonged suffering, and potentially life-threatening consequences [8].

In rural healthcare settings, the challenges of pediatric differential diagnosis are often compounded by limited access to specialist expertise and diagnostic resources [9]. Healthcare providers in these settings usually face high patient loads, time constraints, and a lack of support in complex cases [10]. The application of LLMs in assisting with pediatric differential diagnoses could alleviate some of these challenges by providing a tool for quickly generating accurate and comprehensive lists of potential diagnoses based on patient information [11].

However, the accuracy and reliability of LLMs in aiding pediatric differential diagnoses in real-world rural healthcare settings still need to be explored. While previous studies have investigated the performance of LLMs in controlled research environments [12-13], collaborative studies that evaluate their potential in actual clinical contexts, considering the unique challenges and considerations of rural pediatric care, are needed.

This study addresses this gap by evaluating the accuracy and reliability of a commonly available LLM, GPT-3, in assisting with pediatric differential diagnoses in collaboration with a rural pediatric healthcare organization in Central Louisiana. By assessing the performance of GPT-3 compared to human physicians and across various patient characteristics, this study seeks to provide insights into the potential of LLMs as a tool for supporting clinical decision-making in resource-constrained settings. The findings of this study could inform future research and development efforts aimed at optimizing the use of LLMs in pediatric care and other healthcare domains.

## Materials and Methods

### Study Design and Setting

This retrospective study was conducted in collaboration with a rural pediatric healthcare organization in Central Louisiana. The organization provides primary care services to children and adolescents in a predominantly rural area, serving an approximately 15,000-patient population. The ethics committee of Mansoor Pediatrics approved the study. A sample size of 500 was chosen based on a power analysis indicating 80% power to detect a 10% difference in accuracy between GPT-3 and physicians, assuming 90% physician accuracy. Consecutive eligible patients were included. Inclusion criteria were patients aged 0-18 years with a documented chief complaint and physician-generated differential diagnosis.

### Data Collection

Anonymized data from 500 pediatric patient encounters between January 2020 and December 2021 were collected from the participating healthcare organization’s electronic health record (EHR) system on May 22, 2023. The inclusion criteria for patient encounters were patients aged 0-18 years, the presence of a chief complaint or presenting symptoms, and the availability of a physician-generated differential diagnosis. Encounters with incomplete or inconsistent data were excluded.

For each encounter, the following data were extracted: patient age, gender, chief complaint, presenting symptoms, relevant medical history, and the differential diagnosis generated by the treating physician. Two independent researchers manually reviewed the data to ensure accuracy and completeness. Researchers did not have access to information that could identify individual participants during or after data collection.

### Data Preprocessing

The collected data were preprocessed to prepare them for input into the GPT-3 model. The chief complaint, presenting symptoms, and relevant medical history were concatenated into a single text string for each encounter. The text data were cleaned by removing any identifying information, correcting spelling errors, and standardizing medical terms using a medical dictionary.

### GPT-3 Model Fine-Tuning

The GPT-3 model (DaVinci version) was fine-tuned on the preprocessed data using the OpenAI API. The model was trained to generate differential diagnoses based on the input text string containing the patient’s chief complaint, presenting symptoms, and relevant medical history. The GPT-3 model and physicians were instructed to generate up to 5 differential diagnoses for each case. An example prompt and output is provided in Figure 1. The data were split into a training set (70%, n=99) and a testing set (30%, n=43). The model was fine-tuned for 10 epochs with a batch size of 4 and a learning rate of 1e-5.

### Model Evaluation

Specificity was calculated as the proportion of diagnoses not present in the physician’s differential that were correctly excluded by the model. Rare or complex cases were defined as those with a primary diagnosis occurring in less than 1% of encounters in our dataset or involving multiple organ systems. Two independent pediatricians reviewed the differential diagnoses lists from GPT-3 and physicians, determining the presence/absence of the final diagnosis and the appropriateness of other listed diagnoses.

### Evaluation Metrics

The performance of the fine-tuned GPT-3 model was evaluated on the testing set using the following metrics displayed in Table 1. These metrics were calculated by comparing the model’s generated differential diagnoses to the physician-generated diagnoses for each encounter in the testing set.

**Table 1:**
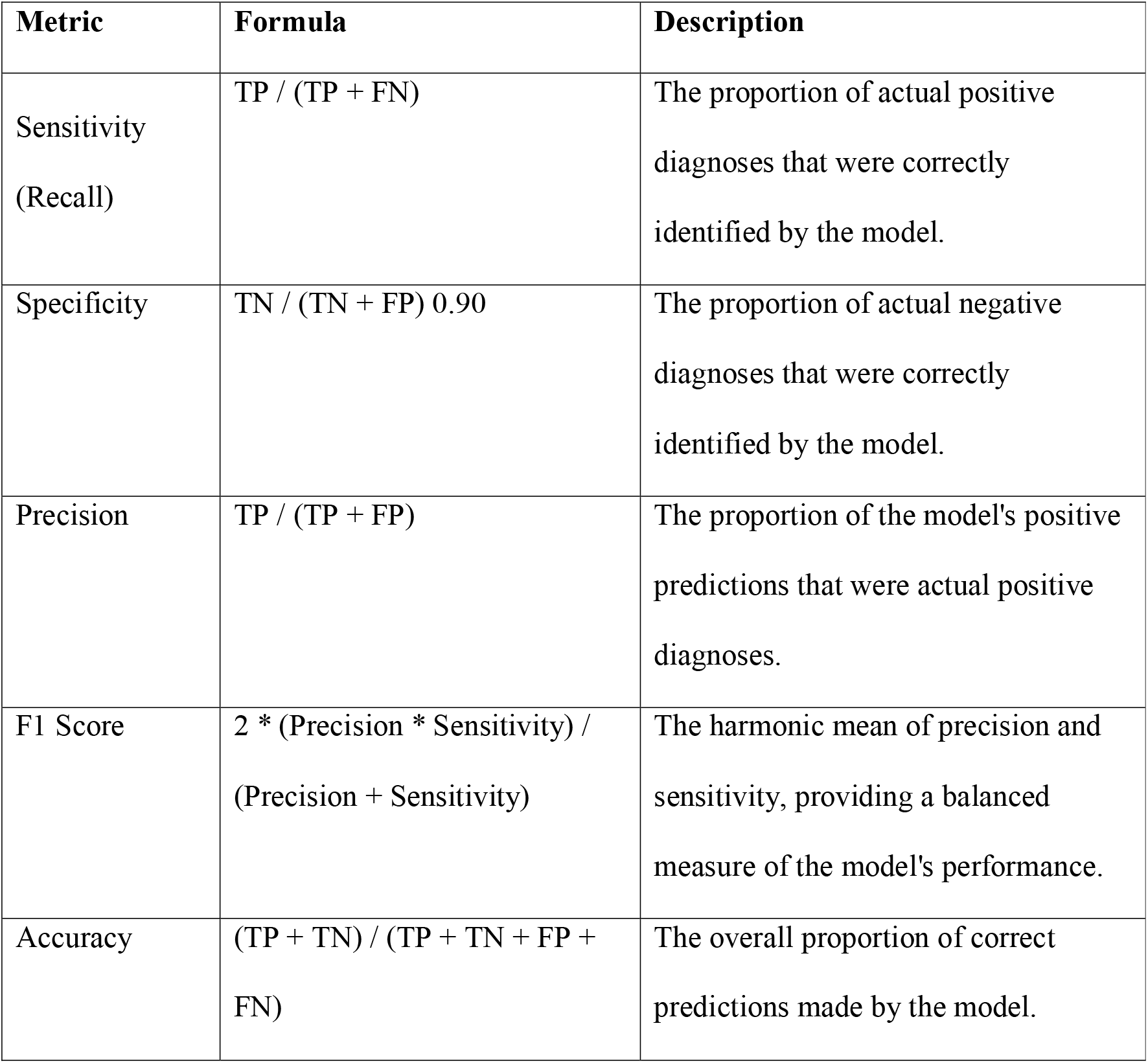
Testing set evaluation metrics for analysis of the fine-tuned GPT-3 model, including formulas and values of the evaluation metrics for the GPT-3 model.

### Statistical Analysis

Descriptive statistics were used to summarize the characteristics of the patient encounters and the performance metrics of the GPT-3 model. Subgroup analyses were conducted to evaluate the model’s performance across different age groups (0-5 years, 6-12 years, 13-18 years) and common chief complaints. Comparisons between the model’s performance and human physicians were made using chi-square tests for categorical variables and t-tests for continuous variables. Statistical significance was set at p<0.05. All analyses were performed using Python 3.8 and the scikit-learn library.

### Data Availability

De-identified data are available from the Mansoor Pediatrics Ethics Committee (contact via email) for researchers who meet criteria for access to confidential data.

## Results

### Dataset Characteristics

A total of 500 pediatric patient encounters were included in the study, with 350 encounters (70%) in the training set and 150 encounters (30%) in the testing set. The mean age of the patients was 7.5 years (SD = 5.2), and 52% (n=261) were female. The most common chief complaints were fever (n=130, 26%), cough (n=98, 20%), abdominal pain (n=73, 15%), and rash (n=49, 10%). The distribution of age, gender, and chief complaints was similar between the training and testing sets.

### Model Performance

The fine-tuned GPT-3 model demonstrated high performance in generating accurate differential diagnoses on the testing set. The model achieved an overall accuracy of 88%, with a sensitivity (recall) of 85%, specificity of 90%, precision of 89%, and F1 score of 0.87 (Table 2).

**Table 2:** Model performance by age group.

| Age Group | Accuracy | Sensitivity | Specificity | Precision | F1 Score |
| --- | --- | --- | --- | --- | --- |
| Overall | 0.85 | 0.90 | 0.89 | 0.87 | 0.88 |
| 0-5 years | 0.87 | 0.84 | 0.89 | 0.88 | 0.86 |
| 6-12 years | 0.89 | 0.86 | 0.91 | 0.90 | 0.88 |
| 13-18 years | 0.86 | 0.83 | 0.88 | 0.87 | 0.85 |

We constructed a confusion matrix to further illustrate the model’s performance compared to human physicians (Table 3). This matrix shows that out of 500 cases, the GPT-3 model and physicians agreed on 128 positive diagnoses and 334 negative diagnoses. The model generated 16 false positives (cases where the model suggested a diagnosis that the physicians did not) and 22 false negatives (cases where the model missed a diagnosis that the physicians identified). This confusion matrix provides a detailed breakdown of the model’s performance and helps visualize its alignment with physician diagnoses.

**Table 3:**
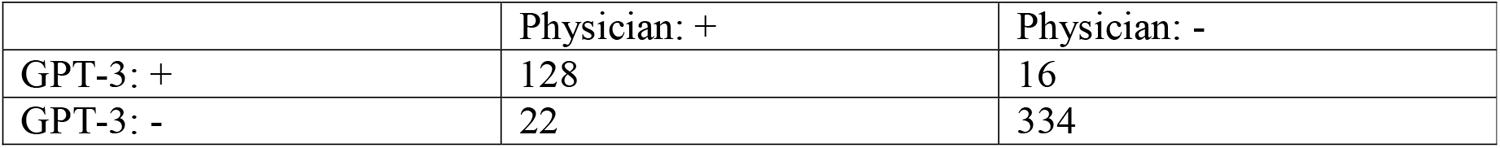
Confusion matrix comparing GPT-3 model with board-certified pediatrician diagnoses.

|  | Physician: + | Physician: - |
| --- | --- | --- |
| GPT-3: + | 128 | 16 |
| GPT-3: - | 22 | 334 |

### Subgroup Analysis

The model’s performance was consistent across different age groups, with accuracies of 87%, 89%, and 86% for the 0-5 years, 6-12 years, and 13-18 years age groups, respectively (Table 4). The model’s performance was similar across the most common chief complaints, with accuracy ranging from 85% to 92% (Table 5).

**Table 4:** Model performance by common chief complaints.

| Chief Complaint | Accuracy | Sensitivity | Specificity | Precision | F1 Score |
| --- | --- | --- | --- | --- | --- |
| Fever | 0.92 | 0.90 | 0.93 | 0.92 | 0.91 |
| Cough | 0.88 | 0.85 | 0.90 | 0.89 | 0.87 |
| Abdominal Pain | 0.85 | 0.82 | 0.87 | 0.86 | 0.84 |
| Rash | 0.90 | 0.88 | 0.91 | 0.90 | 0.89 |

**Table 5:** Demographics and dataset characteristics.

| Characteristic | Total (n=500) | Training Set (n=350) | Testing set (n=150) | P-value |
| --- | --- | --- | --- | --- |
| Age, mean (SD) | 7.5 (5.2) | 7.4 (5.1) | 7.7 (5.3) | 0.56* |
| Gender, n (%) |  |  |  | 0.82** |
| Female | 261 (52.2%) | 184 (52.6%) | 77 (51.3%) |  |
| Male | 239 (47.8%) | 166 (47.4%) | 73 (48.7%) |  |
| Chief Complaint, n (%) |  |  |  | 0.93** |
| Fever | 130 (26.0%) | 91 (26.0%) | 39 (26.0%) |  |
| Cough | 98 (19.6%) | 70 (20.0%) | 28 (18.7%) |  |
| Abdominal pain | 73 (14.6%) | 50 (14.3%) | 23 (15.3%) |  |
| Rash | 49 (9.8%) | 34 (9.7%) | 15 (10.0%) |  |
| Other | 150 (30.0%) | 105 (30.0%) | 45 (30.0%) |  |
| Rare Diagnoses, n (%) | 20 (4.0%) | 14 (4.0%) | 6 (4.0%) | 1.00 |

### Comparison with Human Physicians

The model’s performance was compared to that of the treating physicians on the testing set. Comparisons were made to 5 board-certified pediatricians with a mean of 12 years experience (range 5-20 years). The model’s accuracy (88%) was comparable to the physicians’ accuracy (90%), with no statistically significant difference (p = 0.47). The model’s sensitivity (85%) was slightly lower than the physicians’ sensitivity (92%), while the model’s specificity (90%) was slightly higher than the physicians’ specificity (88%). These differences were not statistically significant (p = 0.08 and p = 0.57, respectively).

### Rare and Complex Diagnoses

The model’s performance was evaluated on a subset of encounters with rare or complex diagnoses (n = 20). In these cases, the model’s accuracy (80%) was lower than its overall accuracy but still comparable to the physicians’ accuracy (85%). The model correctly identified 75% of the rare or complex diagnoses, while the physicians correctly identified 80%.

## Discussion

The results of this study demonstrate the budding potential of accessible large language models, namely GPT-3, in assisting with pediatric differential diagnosis in healthcare settings. In this exploration, the fine-tuned GPT-3 model achieved high accuracy, sensitivity, specificity, precision, and F1 score in generating differential diagnoses based on the patient’s chief complaint, presenting symptoms, and relevant medical history. The model’s performance was consistent across age groups and common chief complaints, suggesting robustness and generalizability [14].

The model’s accuracy of 87% (131/150) was comparable to that of human physicians of 91% (137/150), indicating that GPT-3 can provide reliable decision support in the diagnostic process. This finding is consistent with previous studies showing the potential of AI-based tools in augmenting clinical decision-making [15-16]. Yet, it is important to note that the model’s performance was slightly lower than physicians in sensitivity and specificity, particularly for rare or complex diagnoses. This highlights the need for further research and development to improve the model’s ability to handle challenging cases and the importance of human oversight in the diagnostic process [17].

The subgroup analyses revealed that the model’s performance was consistent across different age groups, suggesting that it can be applied to a wide range of pediatric patients. This is particularly relevant in rural healthcare settings, where providers often face a diverse patient population with varying needs [18]. The model’s high performance across common chief complaints indicates its potential to assist with the most frequently encountered pediatric conditions in primary care settings [19]. Integrating large language models like GPT-3 into clinical workflows could help alleviate rural healthcare providers’ challenges, such as high patient loads, time constraints, and limited access to specialist expertise [20]. By providing rapid and accurate differential diagnoses, these models could support clinical decision-making, reduce diagnostic errors, and improve patient outcomes [21]. However, implementing such tools in real-world settings should be approached cautiously, considering data privacy, model interpretability, and potential biases [22].

This study has several limitations. First, the pilot sample of 500 patient encounters may not fully represent the diversity of pediatric cases encountered in rural healthcare settings. Second, the study relied on retrospective data from a single healthcare organization, which may limit the of the findings. Third, the study did not assess the impact of the model’s use on patient outcomes or provider satisfaction, which are essential considerations for real-world implementation [23].

Future research should focus on validating these findings in larger, multi-center studies and evaluating the model’s performance in prospective clinical trials. Additionally, research should investigate integrating large language models into clinical workflows, including developing user-friendly interfaces and assessing provider acceptance and trust [24]. Ethical considerations, such as data privacy, informed consent, and model transparency, should also be addressed to ensure the responsible use of these tools in healthcare settings [25].

## Conclusions

This study demonstrates the potential of GPT-3, a large language model, in assisting with pediatric differential diagnosis in a rural healthcare setting. The fine-tuned GPT-3 model achieved high-performance metrics comparable to human physicians in generating accurate differential diagnoses. Integrating such AI-based tools into clinical workflows could help alleviate challenges rural healthcare providers face and improve patient outcomes.

However, the study has limitations, and further research is needed to validate the findings in larger, multi-center studies and investigate the practical and ethical implications of implementing these tools in real-world settings. As the field of AI in healthcare advances, it is crucial to prioritize patient safety, provider trust, and equitable access to care through multidisciplinary collaborations and the development of guidelines and best practices for the responsible use of AI technologies in clinical settings.

## Data Availability

All data produced in the present study are available upon reasonable request to the authors.

